# A novel variant of interest of SARS-CoV-2 with multiple spike mutations detected through travel surveillance in Africa

**DOI:** 10.1101/2021.03.30.21254323

**Authors:** Tulio de Oliveira, Silvia Lutucuta, John Nkengasong, Joana Morais, Joana Paula Paixão, Zoraima Neto, Pedro Afonso, Julio Miranda, Kumbelembe David, Luzia Inglês, Amilton Pereira Agostinho Paulo Raisa Rivas Carralero, Helga Reis Freitas, Franco Mufinda, Sofonias Kifle Tessema, Houriiyah Tegally, Emmanuel James San, Eduan Wilkinson, Jennifer Giandhari, Sureshnee Pillay, Marta Giovanetti, Yeshnee Naidoo, Aris Katzourakis, Mahan Ghafari, Lavanya Singh, Derek Tshiabuila, Darren Martin, Richard J Lessells

## Abstract

At the end of 2020, the Network for Genomic Surveillance in South Africa (NGS-SA) detected a SARS-CoV-2 variant of concern (VOC) in South Africa (501Y.V2 or PANGO lineage B.1.351)1. 501Y.V2 is associated with increased transmissibility and resistance to neutralizing antibodies elicited by natural infection and vaccination2,3. 501Y.V2 has since spread to over 50 countries around the world and has contributed to a significant resurgence of the epidemic in southern Africa. In order to rapidly characterize the spread of this and other emerging VOCs and variants of interest (VOIs), NGS-SA partnered with the Africa Centres for Disease Control and Prevention and the African Society of Laboratory Medicine through the Africa Pathogen Genomics Initiative to strengthen SARS-CoV-2 genomic surveillance across the region.

---

Here, we report the first genomic surveillance results from Angola, which has had 21 500 reported cases and around 500 deaths from COVID-19 up to March 2021 (Supplemental Fig S1). On 15 January 2021, in response to the international spread of VOCs, the government instituted compulsory rapid antigen testing of all passengers arriving at the main international airport, in addition to the existing requirement to present a negative PCR test taken within 72 hours of travel. All individuals with a positive antigen test are isolated in a government facility for a minimum of 14 days and require two negative RT-PCR tests at least 48 hours apart for de-isolation, whilst all travelers with a negative test on arrival proceed to mandatory self-quarantine for 10 days followed by a repeat test.

In March 2021, we received 118 nasopharyngeal swab samples collected between June 2020 and February 2021, a number of which were from incoming air travelers (Supplemental Fig S1). From these, we produced 73 high quality genomes (>80% coverage), 14 of which were known VOCs/VOIs (seven 501Y.V2/B.1.351, six B.1.1.7, one B.1.525), 44 of which were C.16 (a common lineage circulating in Portugal), and twelve of which were other lineages (Supplemental Fig S2). In addition, we detected a new VOI in three incoming travelers from Tanzania who were tested together at the airport in mid-February. The three genomes from these passengers were almost identical and presented highly divergent sequences within the A lineage (Figure 1A & 1B). The GISAID database contains nine other sequences reported to be sampled from cases involving travel from Tanzania, two of which are basal to the three sampled in Angola (Figure 1A, Supplemental Table S1).

**Figure 1:**
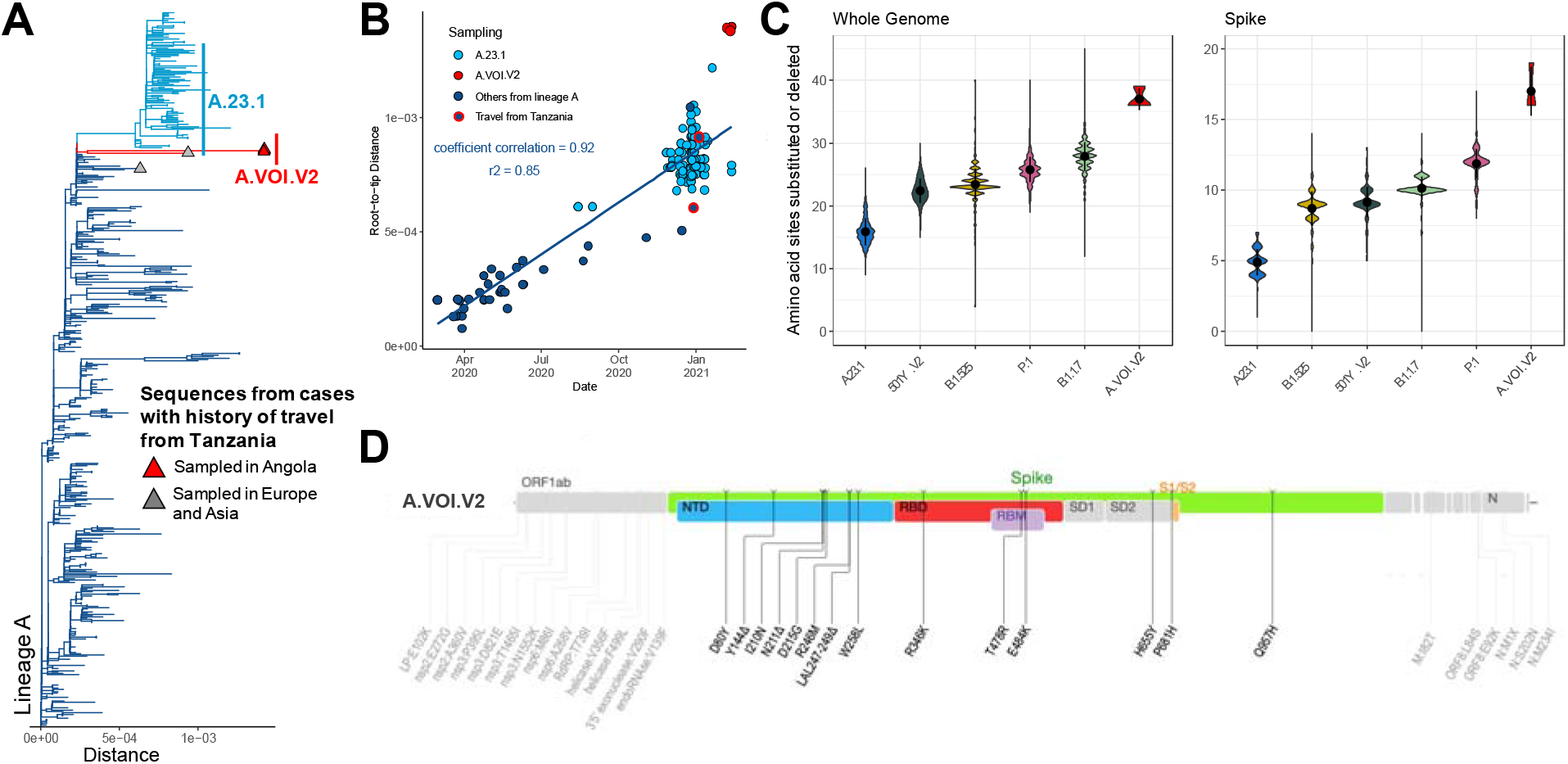
A) Phylogenetic tree of a subset of lineage A sequences (n=319) including five sequences from cases with history of travel of Tanzania, three of which are the A.VOI.V2 sampled in Angola (tips shown with a triangle); B) Regression of root-to-tip genetic distances against sampling dates, for sequences belonging to lineage A, showing the novel A.VOI.V2 (red), the known VOI A.23.1 (light blue), other sequences of lineage A (deep blue), two of which are documented to have travel history from Tanzania (red outline); C) Violin plot showing the number of amino acid mutations in the whole genome and spike glycoprotein in a subset of genomes from five known variants compared to the novel A.VOI.V2; D) Genome map showing the position of the 31 amino acid substitutions and three deletions (spike in color, NTD = N-terminal domain, RBD = receptor-binding domain, RBM = receptor-binding motif, S1/S2 = S1/S2 cleavage site, and the rest of the genome in grey).

This new VOI, temporarily designated A.VOI.V2, has 31 amino acid substitutions (11 in spike) and three deletions (all in spike) (Figure 1C & 1D). The spike mutations include three substitutions in the receptor-binding domain (R346K, T478R and E484K); five substitutions and three deletions in the N-terminal domain, some of which are within the antigenic supersite (Y144&#916;, R246M, SYL247-249&#916; and W258L)4; and two substitutions adjacent to the S1/S2 cleavage site (H655Y and P681H). Several of these mutations are present in other VOCs/VOIs and are evolving under positive selection.

Main Body of the paper (may be some repetion from abstract, due to journal format).

At the end of 2020, the Network for Genomic Surveillance in South Africa (NGS-SA) detected a SARS-CoV-2 variant of concern (VOC) in South Africa (501Y.V2 or PANGO lineage B.1.351)^1^. 501Y.V2 is associated with increased transmissibility and resistance to neutralizing antibodies elicited by natural infection and vaccination^2,3^. 501Y.V2 has since spread to over 50 countries around the world and has contributed to a significant resurgence of the epidemic in southern Africa. In order to rapidly characterize the spread of this and other emerging VOCs and variants of interest (VOIs), NGS-SA partnered with the Africa Centres for Disease Control and Prevention and the African Society of Laboratory Medicine through the Africa Pathogen Genomics Initiative to strengthen SARS-CoV-2 genomic surveillance across the region.

Here, we report the first genomic surveillance results from Angola, which has had 21 500 reported cases and around 500 deaths from COVID-19 up to March 2021 (Supplemental Fig S1). On 15 January 2021, in response to the international spread of VOCs, the government instituted compulsory rapid antigen testing of all passengers arriving at the main international airport, in addition to the existing requirement to present a negative PCR test taken within 72 hours of travel. All individuals with a positive antigen test are isolated in a government facility for a minimum of 14 days and require two negative RT-PCR tests at least 48 hours apart for deisolation, whilst all travelers with a negative test on arrival proceed to mandatory self-quarantine for 10 days followed by a repeat test.

In March 2021, we received 118 nasopharyngeal swab samples collected between June 2020 and February 2021, a number of which were from incoming air travelers (Supplemental Fig S1). From these, we produced 73 high quality genomes (>80% coverage), 14 of which were known VOCs/VOIs (seven 501Y.V2/B.1.351, six B.1.1.7, one B.1.525), 44 of which were C.16 (a common lineage circulating in Portugal), and twelve of which were other lineages (Supplemental Fig S2). In addition, we detected a new VOI in three incoming travelers from Tanzania who were tested together at the airport in mid-February. The three genomes from these passengers were almost identical and presented highly divergent sequences within the A lineage (Figure 1A & 1B). The GISAID database contains nine other sequences reported to be sampled from cases involving travel from Tanzania, two of which are basal to the three sampled in Angola (Figure 1A, Supplemental Table S1).

This new VOI, temporarily designated A.VOI.V2, has 31 amino acid substitutions (11 in spike) and three deletions (all in spike) (Figure 1C & 1D). The spike mutations include three substitutions in the receptor-binding domain (R346K, T478R and E484K); five substitutions and three deletions in the N-terminal domain, some of which are within the antigenic supersite (Y144Δ, R246M, SYL247-249Δ and W258L)^4^; and two substitutions adjacent to the S1/S2 cleavage site (H655Y and P681H). Several of these mutations are present in other VOCs/VOIs and are evolving under positive selection (Figure 1D and Supplemental Text, Fig S2)^5^.

We decided to report this as a new VOI given the constellation of mutations with known or suspected biological significance, specifically resistance to neutralizing antibodies and potentially increased transmissibility (Supplemental Table S3). Whilst we have only detected three cases with this new VOI, this warrants urgent investigation as the source country has a largely undocumented epidemic and few public health measures in place to prevent spread within and out of the country.

<u>Angola Ministry of Health authors:</u>

Silvia Lutucuta (Lutucuta, S)

Zoraima Neto (Neto, Z.)

Pedro Afonso (Afonso, P.)

Julio Miranda (Miranda, J.)

Kumbelembe David (David, K.)

Luzia Inglês (Inglês, L.)

Amilton Pereira (Pereira, A.)

Agostinho Paulo (Paulo, A.)

Raisa Rivas Carralero (Carralero, R. R.)

Joana Paula Paixão (Paixão. J. P.)

Helga Reis Freitas (Freitas R. H.)

Franco Mufinda (Mufinda M.)

Joana Morais (Morais, J.)

<u>AFRICA CDC authors:</u>

John N. Nkengasong (Nkengasong, J. N.)

Sofonias Kifle Tessema (Tessema, S.K.)

<u>KRISP at UKZN authors:</u>

Houriiyah Tegally (Tegally, H.)

Emmanuel James San (San, E.J.)

Eduan Wilkinson (Wilkinson, E.)

Jennifer Giandhari (Giandhari, J.)

Sureshnee Pillay (Pillay, S.)

Yeshnee Naidoo (Naidoo, Y.)

Lavanya Singh (Singh, L.)

Derek Tshiabuila (Tshiabuila, D.)

Richard J. Lessells (Lessells, R. K.)

<u>Fiocruz author:</u>

Marta Giovanetti (Giovanetti, M.)

<u>University of Oxford authors:</u>

Aris Katzourakis (Katzourakis, A.)

Mahan Ghafari (Ghafari, M.)

<u>University of Cape Town author</u>

Darren Martin (Martin, D.)

## Data Availability

All of the sequences are available at GISAID. All of the short reads are available at the Short Read Archive (SRA) of the National Centre for Biotechnology Information (NCBI) (Bioproject Accession: PRJNA717113).

https://www.krisp.org.za

## Notes

### Competing Interest Statement

The authors have declared no competing interest.

### Funding Statement

Funding for genomics surveillance was provided by the South African Medical Research Council (SAMRC), the South African Department of Science and Innovation (DSI), the Africa Centres for Disease Control and Prevention (Africa CDC) and the African Society of Laboratory Medicine (ASLM) through the Africa Pathogen Genomics Initiative (Africa PGI)

### Author Declarations

he project was approved by University of KwaZulu-Natal Biomedical Research Ethics Committee. Protocol reference number: BREC/00001195/2020. Project title: COVID-19 transmission and natural history in KwaZulu-Natal, South Africa: Epidemiological Investigation to Guide Prevention and Clinical Care. Patient consent was not required for the genomic surveillance. This requirement was waived by the Research Ethics Committees.

