## Supplementary Information for "A novel variant of interest of SARS-CoV-2 with multiple spike mutations detected through travel surveillance in Africa"

**SUPPLEMENTAL TEXT**

**Selection analysis**

To identify which, if any, of the observed mutations in the spike protein was most likely to increase viral fitness, we used the natural selection analysis of SARS-CoV-2 pipeline (https://observablehq.com/@

spond/revised-sars-cov-2-analytics-page). This pipeline examines the entire global SARS-CoV-2 nucleotide sequence dataset for evidence of: (i) polymorphisms having arisen in multiple epidemiologically unlinked lineages that have statistical support for non-neutral evolution (Mixed Effects Model of Evolution, MEME)^1^, (ii) sites where these polymorphisms have support for a greater than expected ratio of non-synonymous:synonymous nucleotide substitution rates on internal branches of the phylogenetic tree (Fixed Effects Likelihood, FEL)^2^, and (iii) whether these polymorphisms have increased in frequency in the regions of the world where they have occurred.

Ten of the 30 codons encoding lineage defining amino acid substitutions in the A.VOI.V2 were detectably evolving under positive selection in the 10-Feb-2021 global SARS-CoV2 dataset (P<0.05 using the MEME and FEL selection detection methods (Supplemental Table S4). In addition to the S/478 mutation, this included all five of the codons in the S-gene encoding amino acid substitutions that converge on the signature mutations of 501Y.V1/B.1.1.7 (P681Y), 501Y.V2/B.1.351 (D80Y, D215G, E484K) or 501Y.V3/P.1 (E494K, E484K) variants of concern. This is very strong evidence that mutations at these codons are adaptive either alone or in combination with one another. The four positively selected codons outside of the S-gene were codons Orf1a/540 ORF1a/2283I, ORF8/84 and N/202.

**Supplemental Table S4: Selection analysis of mutated sites on the A.VOI.V2 variant**

| **Gene/Protein** | **Amino Acid Change** | **MEME P- val** | **FEL P- val** | **Date first detected** |
| --- | --- | --- | --- | --- |
| LP | E102K | - | - | - |
| nsp2 | E272G | - | - | - |
| nsp2 | A360V | 0.001 | 0.00056 | 13-May-2020 |
| nsp3 | P395L | - | - | - |
| nsp3 | D821E | - | - | - |
| nsp3 | T1465I | 0.0012 | 0.00061 | 22-Dec-2020 |
| nsp3 | N1552K | - | - | - |
| nsp6 | M86I | - | - | - |
| nsp6 | A268V | - | - | - |
| RdRP | T739I | - | - | - |
| helicase | V356F | - | - | - |
| helicase | F499L | - | - | - |
| 3'5'exonuclease | V290F | - | - | - |
| endoRNAse | V139F | - | - | - |
| S | D80Y | 0.032 | 0.021 | 11-Dec-2020 |
| S | Del Y144 | - | - | - |
| S | I210N | - | - | - |
| S | Del N211 | - | - | - |
| S | D215G | 0.04 | 0.027 | 11-Dec-2020 |
| S | R246M | - | - | - |
| S | Del 247-249 | - | - | - |
| S | W258L | - | - | - |
| S | R346K | - | - | - |
| S | T478R | 0.067 | 0.049 | 10-Feb-2021 |
| S | E484K | 0.013 | 0.0081 | 11-Dec-2020 |
| S | H655Y | 6.30E-05 | 2.70E-05 | 12-Oct-2020 |
| S | P681H | 1.00E-05 | 4.00E-06 | 11-Dec-2020 |
| S | Q957H | - | - | - |
| M | I82T | - | - | - |
| ORF8 | L84S | 0.064 | 0.046 | 31-Mar-2020 |
| ORF8 | E92K | - | - | - |
| N | S202N | 0.0065 | 0.0037 | 13-May-2020 |

**SUPPLEMENTAL METHODS**

Ethical statement

We obtained deidentified remnant nasopharyngeal and oropharyngeal swab samples from patients testing positive for SARS-CoV-2 by RT-qPCR. The project was approved by University of KwaZulu-Natal Biomedical Research Ethics Committee. Protocol reference number: BREC/00001195/2020. Project title: COVID-19 transmission and natural history in KwaZulu-Natal, South Africa: Epidemiological Investigation to Guide Prevention and Clinical Care. Patient consent was not required for the genomic surveillance. This requirement was waived by the Research Ethics Committees.

Epidemiological data

We analyzed COVID-19 cases counts in Angola from publicly released data up to 5^th^ March 2021 from the Our World in Data COVID-19 database (https://github.com/owid/covid-19-data). The estimations for effective daily reproduction number, Re, of SARS-CoV-2 in Angola were obtained from the covid-19-re data repository^3^ (<https://github.com/covid-19-Re/dailyRe-Data>) as at 5^th^ March 2021.

SARS-CoV-2 samples and metadata

Residual samples from nasopharyngeal and oropharyngeal swabs collected from COVID-19 positive patients obtained from Luanda, Angola, were used for SARS-CoV-2 WGS. We obtained samples either in the form of primary swabs or extracted RNA. The swab samples were heat inactivated in a water bath at 60°C for 30 minutes, in biosafety level 3 laboratory, prior to RNA extraction. RNA was extracted using the Viral NA/gDNA Kit on the Chemagic 360 system (Perkin Elmer, Hamburg, Germany) using the automated Chemagic 360 insturment (Perkin Elmer, Hamburg, Germany) or manually using the Qiagen Viral RNA Mini Kit (QIAGEN, California, USA). Associated metadata for the samples included date and location (district) of sampling, and sex and age of the patients, and whether they were community or travel-quarantine cases.

Real Time RT-PCR

In order to detect the SARS-CoV-2 virus by PCR, the TaqPath COVID-19 CE-IVD RT-PCR Kit (Life Technologies, Carlsbad, CA) was used according to the manufacturer’s instructions. The assays target genomic regions (ORF1ab, S protein and N protein) of the SARS-CoV-2 genome. RT-PCR was performed on a QuantStudio 7 Flex Real-Time PCR instrument (Life Technologies, Carlsbad, CA). Cycle thresholds (Ct) values were analyzed using auto-analysis settings with the threshold lines falling within the exponential phase of the fluorescence curves and above any background signal.

Whole genome sequencing and genome assembly

cDNA synthesis was performed on the RNA using random primers followed by gene specific multiplex PCR using the ARTIC protocol^4^. Briefly, extracted RNA was converted to cDNA using the Superscript IV First Strand synthesis system (Life Technologies, Carlsbad, CA) and random hexamer primers. SARS-CoV-2 whole genome amplification by multiplex PCR was carried out using primers designed on Primal Scheme (http://primal.zibraproject.org/) to generate 400bp amplicons with an overlap of 70bp that covers the 30Kb SARS-CoV-2 genome. PCR products were cleaned up using AmpureXP purification beads (Beckman Coulter, High Wycombe, UK) and quantified using the Qubit dsDNA High Sensitivity assay on the Qubit 4.0 instrument (Life Technologies Carlsbad, CA). The Illumina® Nextera Flex DNA Library Prep kit was used according to the manufacturer’s protocol to prepare uniquely indexed paired end libraries of genomic DNA. Sequencing libraries were normalized to 4nM, pooled and denatured with 0.2N sodium acetate. 12pM sample library was spiked with 1% PhiX (PhiX Control v3 adapter-ligated library used as a control). Libraries were loaded onto a 500-cycle v2 MiSeq Reagent Kit and run on the Illumina MiSeq instrument (Illumina, San Diego, CA, USA).

Raw reads coming from Illumina sequencing were assembled using Genome Detective 1.132 (<https://www.genomedetective.com/>) and the Coronavirus Typing Tool^5,6^. All of the sequences were deposited in GISAID (<https://www.gisaid.org/>), and the GISAID accession included as part of the Supplementary Table S1. All raw reads have been deposited to the NCBI Sequence Read Archive (Bioproject Accession: PRJNA717113).

Phylogenetic analysis

Angola sequences in this study were analyzed against a subset of globally representative SARS-CoV-2 genomes, including all genotypes from neighbouring countries, all genotypes in GISAID with known travel history to Tanzania, and enriching for A lineage references. The Angolan sequences were compared against this reference set using a slightly modified version of the SARS-CoV-2 NextStrain build (https://github.com/nextstrain/ncov)^7^. The pipeline contains several python scripts that manage the analysis workflow. In short it allows for the filtering of genotypes, the alignment of genotypes in MAFFT^8^, phylogenetic tree inference in IQ-Tree^9^, tree dating and ancestral state construction and annotation. The lineage A subset of the resulting ML tree was visualized using R ggtree^10^.

Lineage & Clade classification

We used the dynamic lineage classification method proposed by Rambault et al.^11^ in this study via the Phylogenetic Assignment of named Global Outbreak LINeages (PANGOLIN) software suite (<https://github.com/hCoV-2019/pangolin>).

Dated phylogenetics

To estimate time-calibrated phylogenies dated from time-stamped genome data, we conducted phylogenetic analysis using the Bayesian software package BEASTv.1.10.4, on the subset of A sequences including 3 sampled in this study (n=319). The ML tree from this subset were inspected in TempEst v1.5.3 for the presence of a temporal (i.e. molecular clock) signal. Linear regression of root-to-tip genetic distances against sampling dates indicated that the SARS-CoV-2 sequences evolve in a relatively strong clock-like manner (r = 0.92). For this analysis we employed the strict molecular clock model, the HKY+I, nucleotide substitution model and the exponential growth coalescent model^12^. We computed MCMC (Markov chain Monte Carlo) triplicate runs of 100 million states each, sampling every 10.000 steps for each data set. Convergence of MCMC chains was checked using Tracer v.1.7.1. Maximum clade credibility trees were summarised from the MCMC samples using TreeAnnotator after discarding 10% as burn-in.

**SUPPLEMENTAL FIGURES**

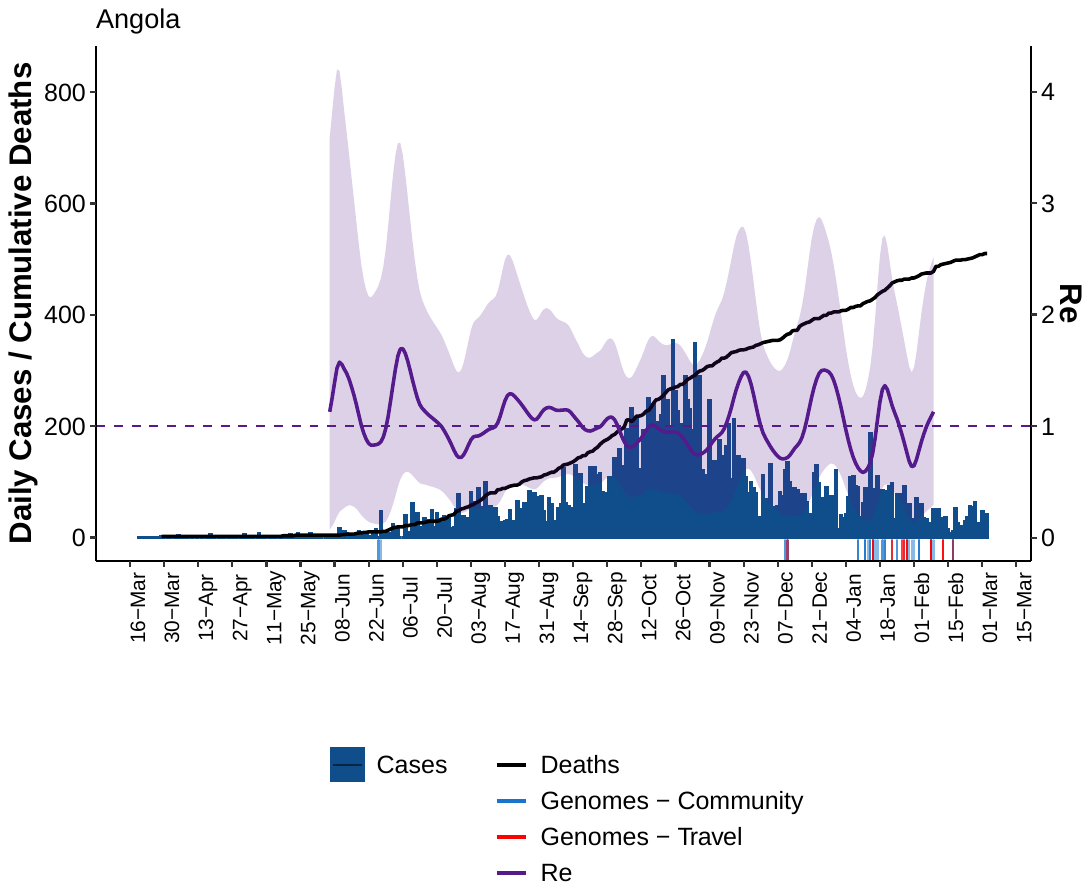

**Supplemental Fig S1:** Epidemiological curve showing the daily recorded case numbers, cumulative deaths and Re estimation in Angola. The rug plot shows the sampling dates of genomes in this study annotated as being either travel cases (red) or community cases (blue).

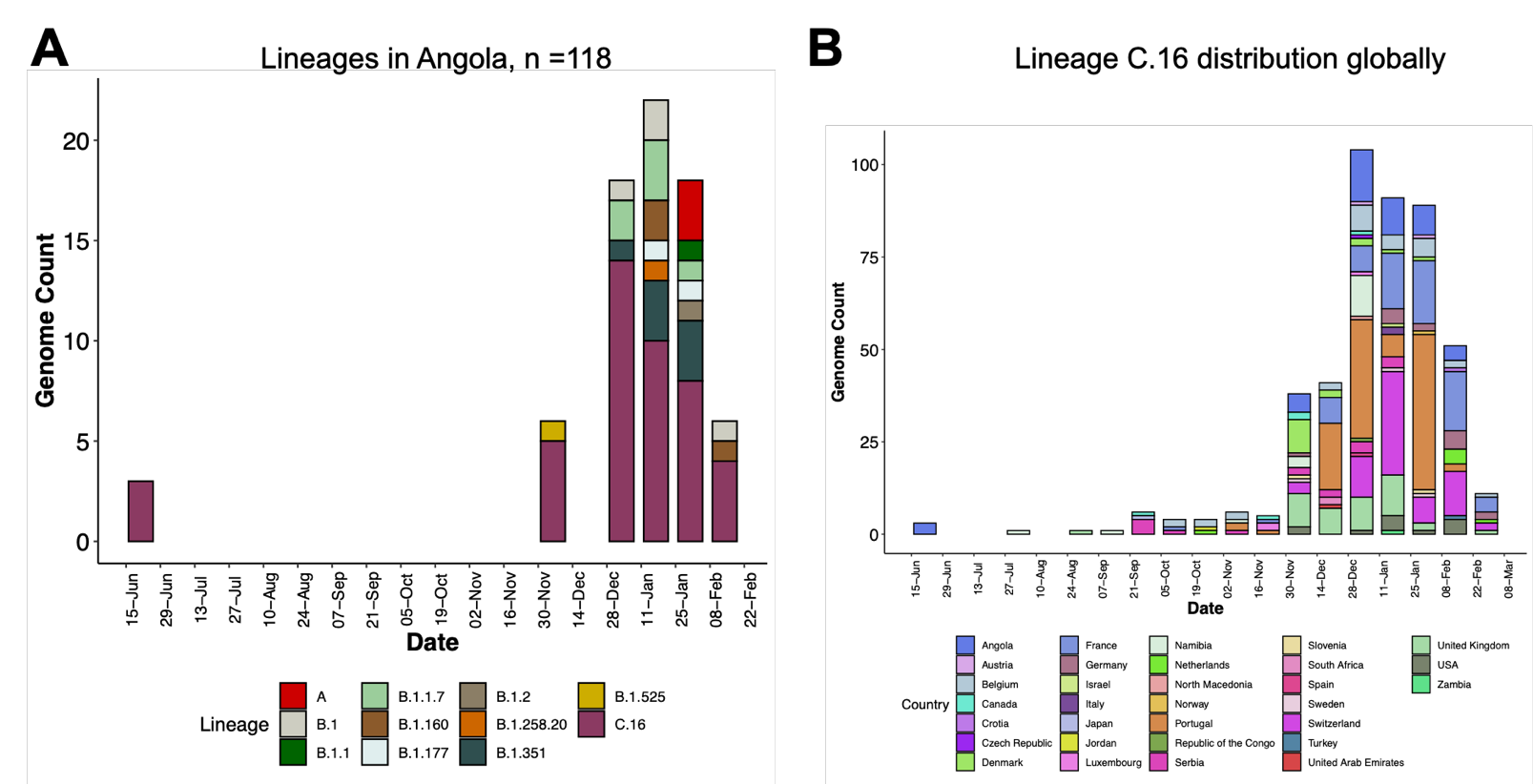

**Supplemental Fig S2: Lineage distribution in Angola.** A) Progression of lineages in Angola by sampling date for all samples sequenced. B) Distribution of the C.16 lineage worldwide against sampling dates.

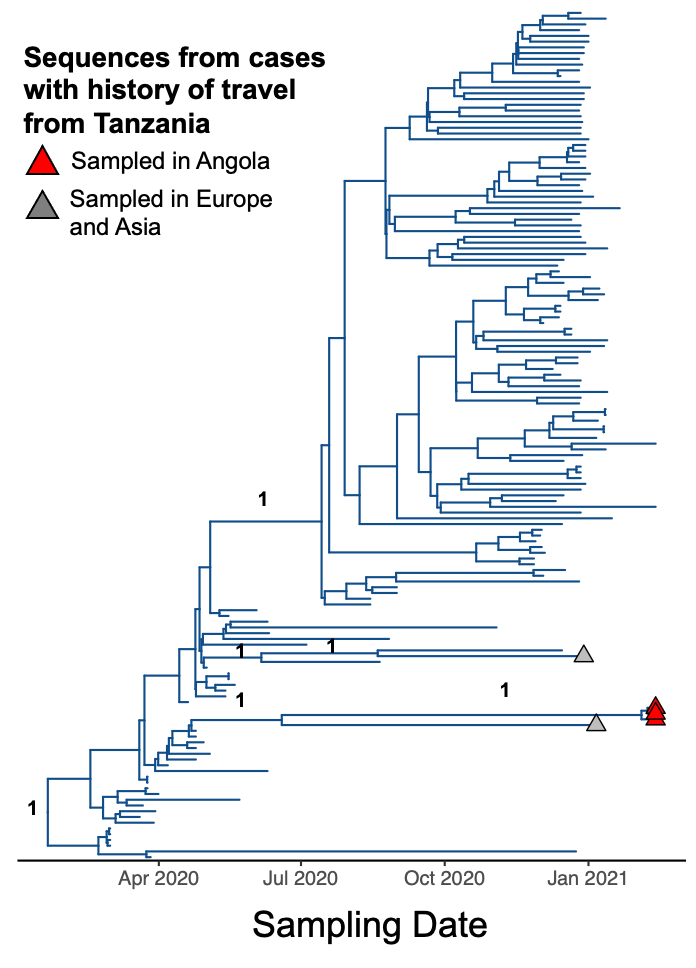

**Supplemental Fig S3:** BEAST tree of A lineage subset (n=319) with selected branch support values shown

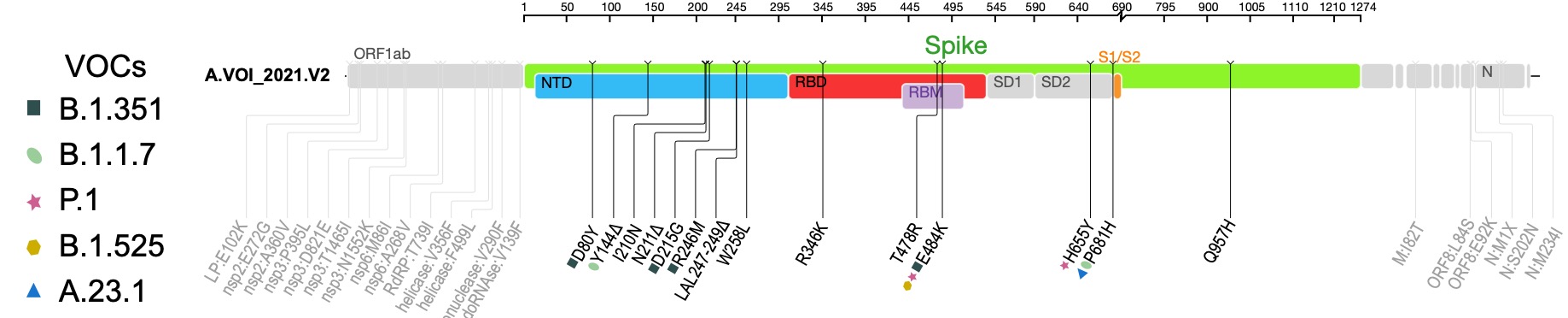

**Supplemental Fig S4:** Genome map of the A.VOI.V2, with mutations annotated by presence in other VOCs

**SUPPLEMENTAL TABLES**

**Supplemental Table S1: Metadata and mutation information for samples associated with travel history from Tanzania with genomes belonging to lineage A.**

| **Sequence Name** | **GISAID Accession** | **Sampling location** | **Sampling date** | **Total Mutations** | **Total Amino Acid Substitutions** | **Nucleotide Substitutions** | **Amino Acid Substitutions** |
| --- | --- | --- | --- | --- | --- | --- | --- |
| **hCoV-19/South Korea/KDCA1491/2020** | **EPI_ISL_1063702** | **South Korea** | 2020/12/30 | 18 | 11 | C337T,G922A,C1190T,C3924T,C8782T,G11230T,C21721T,A23403G,G26389T,T27384C,T27614A,C28000T,G28048T,T28144C,G28167A,G28878A,C29686T,G29742A | E:V49L,N:S202N,ORF1a:P309S,ORF1a:P1220L,ORF1a:M3655I,ORF7a:V74D,ORF8:P36L,ORF8:R52I,ORF8:L84S,ORF8:E92K,S:D614G |
| **hCoV-19/Czech Republic/NRL_277/2021** | **EPI_ISL_882953** | **Czech Republic** | 2021/01/06 | 27 | 23 | T1847C,G2528A,C5907T,T8592C,C8782T,G9273A,C9870T,G10157A,G11230T,T12835C,C15141T,A17423T,A17713C,T19092C,C22916A,C23525T,C24378T,C25613T,C26607T,T26767C,T28144C,G28167A,C28311T,G28878A,G28883A,C29353T,G29742A | M:L29F,M:I82T,N:M1X,N:P13L,N:S202N,N:G204R,ORF1a:S528P,ORF1a:E755K,ORF1a:T1881I,ORF1a:V2776A,ORF1a:R3003K,ORF1a:T3202M,ORF1a:V3298I,ORF1a:M3655I,ORF1b:Y1319F,ORF1b:I1416L,ORF3a:S74F,ORF8:L84S,ORF8:E92K,ORF9b:P10S,S:L452M,S:H655Y,S:S939F |
| **K009791** | **EPI_ISL_1347941** | **Angola** | 2021/02/13 | 44 | 34 | C190T,G569A,T649C,A1620G,C1884T,C3903T,T5182G,C7113T,T7375G,C8782T,G11230T,C11775T,C15279T,C15656T,G17302T,T17731C,G18907T,T20322C,G21073T,G21800T,A22206G,G22335T,A22351G,G22599A,A22742G,A22743C,G22745T,C22995G,G23012A,T23317A,C23525T,C23604A,C23683T,A24433T,T25081C,T26767C,A27198G,G27390C,T28144C,G28167A,G28878A,G28882A,G28975T,G29742A | M:I82T,N:M1X,N:S202N,N:M234I,ORF1a:E102K,ORF1a:E452G,ORF1a:A540V,ORF1a:P1213L,ORF1a:D1639E,ORF1a:T2283I,ORF1a:N2370K,ORF1a:M3655I,ORF1a:A3837V,ORF1b:T730I,ORF1b:V1279F,ORF1b:F1422L,ORF1b:V1814F,ORF1b:V2536F,ORF8:L84S,ORF8:E92K,S:D80Y,S:I210N,S:D215G,S:R246M,S:W258L,S:R346K,S:T393X,S:N394A,S:V395F,S:T478R,S:E484K,S:H655Y,S:P681H,S:Q957H |
| **K009790** | **EPI_ISL_1347940** | **Angola** | 2021/02/13 | 41 | 31 | C190T,G569A,T649C,A1620G,C1884T,C3903T,T5182G,C7113T,T7375G,C8782T,G11230T,C11775T,C15279T,C15656T,G17302T,T17731C,G18907T,T20322C,G21073T,G21800T,A22206G,G22335T,A22351G,G22599A,C22995G,G23012A,T23317A,C23525T,C23604A,C23683T,A24433T,T25081C,T26767C,A27198G,G27390C,T28144C,G28167A,G28878A,G28882A,G28975T,G29742A | M:I82T,N:M1X,N:S202N,N:M234I,ORF1a:E102K,ORF1a:E452G,ORF1a:A540V,ORF1a:P1213L,ORF1a:D1639E,ORF1a:T2283I,ORF1a:N2370K,ORF1a:M3655I,ORF1a:A3837V,ORF1b:T730I,ORF1b:V1279F,ORF1b:F1422L,ORF1b:V1814F,ORF1b:V2536F,ORF8:L84S,ORF8:E92K,S:D80Y,S:I210N,S:D215G,S:R246M,S:W258L,S:R346K,S:T478R,S:E484K,S:H655Y,S:P681H,S:Q957H |
| **K009792** | **EPI_ISL_1347942** | **Angola** | 2021/02/13 | 41 | 31 | C190T,G569A,T649C,A1620G,C1884T,C3903T,T5182G,C7113T,T7375G,C8782T,G11230T,C11775T,C15279T,C15656T,G17302T,T17731C,G18907T,T20322C,G21073T,G21800T,A22206G,G22335T,A22351G,G22599A,C22995G,G23012A,T23317A,C23525T,C23604A,C23683T,A24433T,T25081C,T26767C,A27198G,G27390C,T28144C,G28167A,G28878A,G28882A,G28975T,G29742A | M:I82T,N:M1X,N:S202N,N:M234I,ORF1a:E102K,ORF1a:E452G,ORF1a:A540V,ORF1a:P1213L,ORF1a:D1639E,ORF1a:T2283I,ORF1a:N2370K,ORF1a:M3655I,ORF1a:A3837V,ORF1b:T730I,ORF1b:V1279F,ORF1b:F1422L,ORF1b:V1814F,ORF1b:V2536F,ORF8:L84S,ORF8:E92K,S:D80Y,S:I210N,S:D215G,S:R246M,S:W258L,S:R346K,S:T478R,S:E484K,S:H655Y,S:P681H,S:Q957H |

**Supplemental Table S2: Putative biological effects of A.VOI.V2 spike mutations**

| **Spike Mutations** | **Spike subregion** | **Present in other VOC/VOI** | **Potential biological effects** | **References** |
| --- | --- | --- | --- | --- |
| D80Y | N-terminal domain | B.1.351 (*D80A*) | Mutations at that position associated with resistance to NTD mAbs | McCallum^13^ |
| Y144Δ | N-terminal domain | B.1.1.7 | Forms part of NTD antigenic supersite; deletion disrupts binding of mAbs; strongly associated with resistance to NTD mAbs | McCallum^13^  McCarthy^14^ |
| I210N | N-terminal domain | - | - | - |
| N211Δ | N-terminal domain | - | - | - |
| D215G | N-terminal domain | B.1.351 | - | - |
| R246M | N-terminal domain | B.1.351 (*R246I*) | Forms part of NTD antigenic supersite; mutations at that residue associated with resistance to NTD mAbs | McCallum^13^ |
| SYL247-249Δ | N-terminal domain | - | Forms part of NTD antigenic supersite; deletions may be associated with resistance to NTD mAbs | McCallum^13^ |
| W258L | N-terminal domain | - | Forms part of NTD antigenic supersite; mutations at that residue may be associated with resistance to NTD mAbs | McCallum^13^ |
| R346K | Receptor-binding domain | - | Key site for binding of class 3 RBD NAbs; mutation associated with resistance to class 3 RBD NAbs | Barnes^15^  Weisblum^16^  Greaney^17^  Liu^18^ |
| T478R | Receptor-binding domain (receptor-binding motif) |  | Mutations at that position associated with resistance to mAbs | Liu^18^ |
| E484K | Receptor-binding domain  (receptor-binding motif) | B.1.351  P.1  P.2  B.1.525 | Key site for binding of class 2 RBD NAbs; mutation associated with resistance to class 2 RBD NAbs and resistance to polyclonal sera | Barnes^15^  Weisblum^16^  Baum^19^  Greaney^17,20,21^  Liu^18^ |
| H655Y | Adjacent to S1/S2 cleavage site | P.1 | - | - |
| P681H | S1/S2 cleavage site | B.1.1.7  A.23.1 (*P681R*) | Mutations may affect efficiency of cell entry | Peacock^22^ |
| Q957H | Heptad-repeat 1 | - | - | - |

**Supplemental Table S3: GISAID Acknowledgements.**

| **strain** | **gisaid_epi_isl** | **originating_lab** | **submitting_lab** | **authors** |
| --- | --- | --- | --- | --- |
| Australia/SAP603/2021 | EPI_ISL_771368 | SA Pathology | SA Pathology | Lex Leong et al |
| Botswana/CV2707886/2021 | EPI_ISL_944767 | Botswana Harvard HIV Reference Laboratory | Botswana Harvard HIV Reference Laboratory | Sikhulile Moyo et al |
| Cambodia/126518/2020 | EPI_ISL_918370 | Virology Unit, Institut Pasteur du Cambodge | Virology Unit, Institut Pasteur du Cambodge | Sokhoun Yann et al |
| Cambodia/126521/2020 | EPI_ISL_918372 |  |  |  |
| Cambodia/127828/2020 | EPI_ISL_918366 |  |  |  |
| Cambodia/137333/2020 | EPI_ISL_918365 |  |  |  |
| Cambodia/137931/2020 | EPI_ISL_918364 |  |  |  |
| Cambodia/137932/2020 | EPI_ISL_918363 |  |  |  |
| Cambodia/139820/2020 | EPI_ISL_918361 |  |  |  |
| CzechRepublic/NRL_277/2021 | EPI_ISL_882953 | The National Institute of Public Health | State Veterinary Institute Prague | Nagy et al |
| Denmark/DCGC-855/2020 | EPI_ISL_614814 | Department of Virus and Microbiological Special Diagnostics, Statens Serum Institut, Denmark | Albertsen lab, Department of Chemistry and Bioscience, Aalborg University, Denmark | Danish Covid-19 Genome Consortia et al |
| DRC/04186/2020 | EPI_ISL_471411 | Viral Respiratory Lab, National Institute for Biomedical Research (INRB) | Pathogen Sequencing Lab, National Institute for Biomedical Research (INRB) | Placide Mbala-Kingebeni et al |
| DRC/04187/2020 | EPI_ISL_471412 |  |  |  |
| DRC/04188/2020 | EPI_ISL_471413 |  |  |  |
| DRC/3834/2020 | EPI_ISL_447247 |  |  |  |
| DRC/4524/2020 | EPI_ISL_513597 |  |  |  |
| Egypt/CUNCI-HGC9I022/2020 | EPI_ISL_857332 | Cancer Biology Department, National Cancer Institute | Cancer Biology Department, National Cancer Institute | Zekri et al |
| Gabon/ITM-K027/2020 | EPI_ISL_539575 | Centre de Recherches Medicales de Lambarene (CERMEL) | Department of Emerging Infectious Diseases, Institute of Tropical Medicine, Nagasaki University | Haruka Abe et al |
| Ghana/1651_S3/2020 | EPI_ISL_422387 | NMIMR, Department of Virology | WACCBIP, University of Ghana | Joyce M. Ngoi et al |
| Ghana/2828_S6/2020 | EPI_ISL_422397 |  |  |  |
| Ghana/3177_S12/2020 | EPI_ISL_422403 |  |  |  |
| Ghana/34927_S20/2020 | EPI_ISL_515086 | Department of Biochemistry, Cell and Molecular Biology | WACCBIP, University of Ghana | Ngoi et al |
| Ghana/KATH23/2020 | EPI_ISL_515181 | Kumasi Centre for Collaborative Research in Tropical Medicine, Kumasi. | Institute of Virology, Charit√© ‚Äì Universit√§tsmedizin Berlin | Augustina Sylverken et al |
| Kenya/C14075/2020 | EPI_ISL_806643 | KEMRI-Wellcome Trust Research Programme/KEMRI-CGMR-C Kilifi | KEMRI-Wellcome Trust Research Programme/KEMRI-CGMR-C Kilifi | Githinji et al et al |
| Kenya/C6519/2020 | EPI_ISL_568804 |  |  |  |
| Kenya/C6524/2020 | EPI_ISL_568805 |  |  |  |
| Kenya/C7579/2020 | EPI_ISL_568842 |  |  |  |
| Kenya/C7580/2020 | EPI_ISL_568843 |  |  |  |
| Kenya/C7605/2020 | EPI_ISL_568847 |  |  |  |
| Kenya/C76749/2020 | EPI_ISL_855544 |  |  |  |
| Kenya/C77606/2020 | EPI_ISL_855546 |  |  |  |
| Kenya/C80125/2021 | EPI_ISL_969003 |  |  |  |
| Kenya/C8439/2020 | EPI_ISL_568863 |  |  |  |
| Mauritius/N2023/2021 | EPI_ISL_1191599 | Virology Department, Victoria Hospital, Plaine-Wilhems, Mauritius | National Institute for Communicable Diseases of the National Health Laboratory Service | Ramuth M et al |
| Nigeria/ED04-CV158/2020 | EPI_ISL_527876 | Nigeria Centre for Disease Control (NCDC) | African Centre of Excellence for Genomics of Infectious Diseases (ACEGID), Redeemer's University, Ede, Osun State, Nigeria | Oluniyi P.E. et al et al |
| Nigeria/ED06-CV159/2020 | EPI_ISL_527877 | Nigeria Centre for Disease Control (NCDC) | African Centre of Excellence for Genomics of Infectious Diseases (ACEGID), Redeemer's University, Ede, Osun State, Nigeria | Oluniyi P.E. et al et al |
| Rwanda/AT33505RD/2020 | EPI_ISL_1063901 | Rwanda National Reference Laboratory | Rwanda National Reference Laboratory | Enatha Mukantwari et al |
| Rwanda/AX81260RD/2020 | EPI_ISL_1064147 |  |  |  |
| Rwanda/AX83834RD/2020 | EPI_ISL_1064148 |  |  |  |
| Rwanda/AY77279RD/2021 | EPI_ISL_1064164 |  |  |  |
| Rwanda/BH72124RD/2021 | EPI_ISL_1064168 |  |  |  |
| Rwanda/BH74409RD/2021 | EPI_ISL_1064170 |  |  |  |
| Rwanda/NRLNAT1010/2020 | EPI_ISL_925848 | Nucleic Acid Testing, National Reference Laboratory | GIGA Medical Genomics | Yvan Butera et al |
| Rwanda/NRLNAT1011/2020 | EPI_ISL_925849 |  |  |  |
| Rwanda/NRLNAT1012/2020 | EPI_ISL_925850 |  |  |  |
| Rwanda/NRLNAT1013/2020 | EPI_ISL_925851 |  |  |  |
| Rwanda/NRLNAT1014/2020 | EPI_ISL_925852 |  |  |  |
| Rwanda/NRLNAT1018/2020 | EPI_ISL_925856 |  |  |  |
| Rwanda/NRLNAT1019/2020 | EPI_ISL_925857 |  |  |  |
| Rwanda/NRLNAT1020/2020 | EPI_ISL_925858 |  |  |  |
| Rwanda/NRLNAT1024/2020 | EPI_ISL_925861 |  |  |  |
| Rwanda/NRLNAT1029/2020 | EPI_ISL_925865 |  |  |  |
| Rwanda/NRLNAT1039/2020 | EPI_ISL_925875 |  |  |  |
| Rwanda/NRLNAT1047/2021 | EPI_ISL_925882 |  |  |  |
| Rwanda/NRLNAT1049/2021 | EPI_ISL_925884 |  |  |  |
| Rwanda/NRLNAT1050/2021 | EPI_ISL_925885 |  |  |  |
| Rwanda/NRLNAT1051/2021 | EPI_ISL_925886 |  |  |  |
| Rwanda/NRLNAT1052/2021 | EPI_ISL_925887 |  |  |  |
| Rwanda/NRLNAT1053/2021 | EPI_ISL_925888 |  |  |  |
| Rwanda/NRLNAT1054/2021 | EPI_ISL_925889 |  |  |  |
| Rwanda/NRLNAT1055/2021 | EPI_ISL_925890 |  |  |  |
| Rwanda/NRLNAT1057/2021 | EPI_ISL_925892 |  |  |  |
| Rwanda/NRLNAT1059/2020 | EPI_ISL_925894 |  |  |  |
| Rwanda/NRLNAT1060/2020 | EPI_ISL_925895 |  |  |  |
| Rwanda/NRLNAT1062/2020 | EPI_ISL_925897 |  |  |  |
| Rwanda/NRLNAT1064/2020 | EPI_ISL_925898 |  |  |  |
| Rwanda/NRLNAT1079/2021 | EPI_ISL_925908 |  |  |  |
| Rwanda/NRLNAT1080/2021 | EPI_ISL_925909 |  |  |  |
| Rwanda/NRLNAT1081/2021 | EPI_ISL_925910 |  |  |  |
| Rwanda/NRLNAT1083/2021 | EPI_ISL_925912 |  |  |  |
| Rwanda/NRLNAT1085/2021 | EPI_ISL_925914 |  |  |  |
| Rwanda/NRLNAT1086/2021 | EPI_ISL_925915 |  |  |  |
| Senegal/1966/2020 | EPI_ISL_480789 | Institut Pasteur Dakar | Institut Pasteur de Dakar | Ndongo Dia et al |
| Senegal/620/2020 | EPI_ISL_420078 |  |  |  |
| Singapore/1390/2020 | EPI_ISL_645117 | National Public Health Laboratory, National Centre for Infectious Diseases | National Public Health Laboratory, National Centre for Infectious Diseases | Tze Minn Mak et al |
| SouthKorea/KDCA1491/2020 | EPI_ISL_1063702 | Division of Emerging Infectious Diseases, Bureau of Infectious Diseases Diagnosis Control, Korea Disease Control and Prevention Agency | Division of Emerging Infectious Diseases, Bureau of Infectious Diseases Diagnosis Control, Korea Disease Control and Prevention Agency | Ae Kyung Park et al |
| Uganda/UG017/2020 | EPI_ISL_451199 | Uganda Virus Research Institute | MRC/UVRI & LSHTM Uganda Research Unit | Dan Lule Bugembe et al |
| Uganda/UG042/2020 | EPI_ISL_737962 | Uganda Central Public Health Lab and Uganda Virus Research Institute | MRC/UVRI & LSHTM Uganda Research Unit | Matthew Cotten et al |
| Uganda/UG115/2020 | EPI_ISL_738022 |  |  |  |
| Uganda/UG125/2020 | EPI_ISL_738032 |  |  |  |
| Uganda/UG128/2020 | EPI_ISL_738035 |  |  |  |
| Uganda/UG163/2020 | EPI_ISL_954238 | MRC/UVRI & LSHTM Uganda Research Unit | | Matthew Cotten et al |
| Uganda/UG164/2020 | EPI_ISL_954239 |  |  |  |
| Uganda/UG165/2020 | EPI_ISL_954255 |  |  |  |
| Uganda/UG166/2020 | EPI_ISL_954229 |  |  |  |
| Uganda/UG167/2020 | EPI_ISL_954261 |  |  |  |
| Uganda/UG168/2020 | EPI_ISL_954262 |  |  |  |
| Uganda/UG169/2020 | EPI_ISL_954263 |  |  |  |
| Uganda/UG170/2020 | EPI_ISL_954264 |  |  |  |
| Uganda/UG171/2020 | EPI_ISL_954266 |  |  |  |
| Uganda/UG172/2020 | EPI_ISL_954267 |  |  |  |
| Uganda/UG173/2020 | EPI_ISL_954256 |  |  |  |
| Uganda/UG174/2020 | EPI_ISL_954257 |  |  |  |
| Uganda/UG175/2020 | EPI_ISL_954258 |  |  |  |
| Uganda/UG176/2020 | EPI_ISL_954259 |  |  |  |
| Uganda/UG177/2020 | EPI_ISL_954260 |  |  |  |
| Uganda/UG178/2020 | EPI_ISL_954268 |  |  |  |
| Uganda/UG179/2020 | EPI_ISL_954269 |  |  |  |
| Uganda/UG180/2020 | EPI_ISL_954227 |  |  |  |
| Uganda/UG181/2020 | EPI_ISL_954270 |  |  |  |
| Uganda/UG182/2020 | EPI_ISL_954271 |  |  |  |
| Uganda/UG183/2020 | EPI_ISL_954272 |  |  |  |
| Uganda/UG184/2020 | EPI_ISL_954282 |  |  |  |
| Uganda/UG185/2020 | EPI_ISL_955136 |  |  |  |
| Uganda/UG186/2020 | EPI_ISL_954283 |  |  |  |
| Uganda/UG187/2020 | EPI_ISL_954281 |  |  |  |
| Uganda/UG188/2020 | EPI_ISL_954284 |  |  |  |
| Uganda/UG189/2020 | EPI_ISL_954288 |  |  |  |
| Uganda/UG190/2020 | EPI_ISL_954289 |  |  |  |
| Uganda/UG191/2020 | EPI_ISL_954285 |  |  |  |
| Uganda/UG192/2020 | EPI_ISL_954286 |  |  |  |
| Uganda/UG193/2020 | EPI_ISL_954287 |  |  |  |
| Uganda/UG194/2020 | EPI_ISL_954290 |  |  |  |
| Uganda/UG195/2020 | EPI_ISL_954291 |  |  |  |
| Uganda/UG196/2020 | EPI_ISL_954292 |  |  |  |
| Uganda/UG197/2020 | EPI_ISL_954293 |  |  |  |
| Uganda/UG198/2020 | EPI_ISL_954294 |  |  |  |
| Uganda/UG199/2020 | EPI_ISL_954295 |  |  |  |
| Uganda/UG200/2020 | EPI_ISL_954296 |  |  |  |
| Uganda/UG201/2021 | EPI_ISL_954297 |  |  |  |
| Uganda/UG202/2021 | EPI_ISL_954298 |  |  |  |
| Uganda/UG203/2021 | EPI_ISL_954299 |  |  |  |
| Uganda/UG204/2021 | EPI_ISL_954300 |  |  |  |
| Uganda/UG205/2021 | EPI_ISL_954226 |  |  |  |
| Uganda/UG206/2020 | EPI_ISL_954273 |  |  |  |
| Uganda/UG207/2020 | EPI_ISL_954274 |  |  |  |
| Uganda/UG208/2020 | EPI_ISL_954275 |  |  |  |
| Uganda/UG209/2020 | EPI_ISL_954276 |  |  |  |
| Uganda/UG210/2020 | EPI_ISL_954277 |  |  |  |
| Uganda/UG211/2020 | EPI_ISL_954278 |  |  |  |
| Uganda/UG213/2020 | EPI_ISL_954279 |  |  |  |
| Uganda/UG214/2020 | EPI_ISL_954280 |  |  |  |
| UnitedArabEmirates/0993/2020 | EPI_ISL_699126 | Group 42 (G42) Healthcare, Abu Dhabi, United Arab Emirates; Department of Health, The United Arab Emirates | G42 Healthcare | Rong Liu et al |
| UnitedArabEmirates/1039/2020 | EPI_ISL_698114 |  |  |  |
| UnitedArabEmirates/1881/2020 | EPI_ISL_859637 | BTC, Khalifa University | BTC, Khalifa University | Al Safar et al et al |
| UnitedArabEmirates/2002/2020 | EPI_ISL_859646 |  |  |  |
| Zambia/ZMB-1241/2020 | EPI_ISL_977357 | University of Zambia, School of Veterinary Medicine | UNZAVET and PATH | Mulenga Mwenda-Chimfwembe et al |
| Zambia/ZMB-23199/2020 | EPI_ISL_977370 |  |  |  |
| Zimbabwe/ZW-149/2020 | EPI_ISL_644750 | National Microbiology Reference Laboratory | Quadram Institute Bioscience | Thanh Le Viet et al |
| Zimbabwe/ZW-168/2020 | EPI_ISL_644743 |  |  |  |
| Zimbabwe/ZW-169/2020 | EPI_ISL_647976 |  |  |  |
| Zimbabwe/ZW-214/2020 | EPI_ISL_644764 |  |  |  |
| Zimbabwe/ZW-41190/2020 | EPI_ISL_1191853 |  |  | Tapfumanei Mashe et al |
| Zimbabwe/ZW-41817/2020 | EPI_ISL_1191887 |  |  |  |
| Zimbabwe/ZW-41820/2020 | EPI_ISL_1191888 |  |  |  |

**ACKNOWLEDGEMENTS**

We would like to thank Robert Shafer for contributing the genome map pipeline, and would like to thank Jessie Bloom, Allie Greaney and Richard Neher for helpful comments on the significance of the mutations identified in the A.VOI.V2 variant.

6.	Cleemput S, Dumon W, Fonseca V, et al. Genome Detective Coronavirus Typing Tool for rapid identification and characterization of novel coronavirus genomes.
